# Adequacy Of Salt Iodization, Retailers’ Knowledge And Associated Factors About Iodized Salt In Oromia Special Zone Surrounding Finfinne, Ethiopia

**DOI:** 10.1101/2022.03.29.22272937

**Authors:** Meron kebede, Zewdie Aderaw Alemu, Dano Gutata

## Abstract

**Background:** There are laws to enforce the universal iodization of salt to check the consequences of iodine deficiency in Ethiopia. These laws are to ensure that there are production and sales of iodized salt in the country. Yet, the adequacy of iodized salt in the retailers’ level is not determined.

**Objective:** This study is aimed to investigate the adequacy of salt iodization, retailers’ knowledge, and associated factors about iodized salt among retailers in Addis Ababa surrounding finfinne Special Zone, Ethiopia.

**Methods:** A community-based cross-sectional study was conducted among 202 iodized salt retailers that were selected by using a simple random sampling technique. Data among the shop retailers for the assessment of knowledge was collected by using a structured questionnaire, and titration test method was done to determine the adequacy of salt iodization. The data management and analysis were done by using Statistical Packages for Social Scientists Version 23. Descriptive statistics such as mean, median, standard deviation, and frequency were computed to describe the data. Binary logistic regression analysis was done to examine the association between the dependent and independent variables. The strength of association between the dependent and independent variables was explained by using the odds ratio and the level of statistical significance was accepted at a p-value of less than 0.05. Finally, the results were presented by using graphs and tables.

**Result:** A total of 202 salt retailers were approached in this study with a mean age of 28.78±7.548 years. Nearly around two-thirds of the study participants were male. Of the total retailers’ salt samples, 95(57.2%) were adequately (20-40ppm) iodized, and the educational level of the retailers was significantly associated with the adequacy of salt iodization. From the total salt retailer, 113(55.9%) had good knowledge, and retailers who heard about iodized salt (AOR=0.043, CI: 0.017-0.109, p<0.001) and having legal framework/law (AOR=0.24, CI: 0.098-0.61, p=0.003) that prohibits the selling of non-iodized salt were significantly associated with good knowledge level.

**Conclusion and Recommendation:** Still policies have been implemented to promote the production and consumption of iodized salt, the iodine content of salt in retail shops in the Oromia Special Zone Finfinne, Oromia regional state is not encouraging. We recommend the establishment of checkpoints along the production and distribution chain to ensure salt with adequate iodine reaches the consumer. Again, traders of iodized salt should have regular training on ways to preserve salt to maintain its iodine content

## Introduction

Iodine is a micronutrient that is needed for the production of thyroid hormone called thyroxin, and the body does not make iodine, as a result, it is an essential part of the diet, which prevents iodine deficiency disorder (IDD), a global public health concern in many developing countries including Ethiopia(1). This micronutrient is a vital element of the thyroid gland for thyroid hormone production, growth, and development of the brain and central nervous system(2,3). A deficiency of iodine leads to low production of thyroid hormones, which have a wide range of negative implications on different organs and muscle functions such as generally heart, liver, kidneys and development of the brain(4), stillbirths, congenital abnormalities, and decreased cognitive capacity(5), and goiter or enlargement of the thyroid gland (3,4).

Iodine is found in various foods such as cheese, cow’s milk, eggs, frozen yogurt, ice cream, iodine-containing multivitamins, iodized table salt, saltwater fish, seaweed, shellfish, soy milk, soy sauce, and yogurt(6).

World health organization (WHO) recommends that the median iodine urinary level need to be within the range (100–199 μg/l) to ensure adequate iodine content in salt and other sources of iodine in the diet or iodized salt containing 20 to 40 ppm of iodine at the household level regarded as adequately iodized(7).

Iodization of salt is the first-line measure to prevent and control iodine deficiency disorders. The Ethiopian Council of Ministers passed salt legislation in March 2011; according to this regulation, every salt for human consumption needs to be iodized, and any iodized salt for human consumption shall conform to the standards for iodized salt set by the appropriate authority(8).

### Universal Salt Iodization (USI), which intends that all salt for human and animal

consumption be iodized thus ensuring adequate iodine nutrition, was identified as the global strategy for the elimination of iodine deficiency. Salt is an excellent carrier for iodine and other nutrients as it is safe, consumed at relatively constant, well-definable levels by all people within a society, independently of economic status(9).

Iodized salt has been credited with preventing about 750 million cases of goiter over the past years, with Iodine Global Network and UNICEF estimating that globally about 6.1 billion people are currently consuming iodized salt representing a significant achievement of large-scale food fortification(10).

Iodine Deficiency (ID) is the most common preventable cause of intellectual impairment. Globally, 241 million populations are estimated to have insufficient iodine intakes. The problem is worse in Southeast Asia and Africa, where 76 million and 58 million populations have inadequate iodine intake respectively (11). According to the Ethiopian National Nutrient survey report, 47.5% of schoolchildren had urinary iodine levels less than 100 μg/L. The median Urinary iodine level in non-pregnant women of reproductive age was 96.8 μg/L; about 51.8% of women had urinary iodine levels less than 100 μg/L. The report also showed that only 26% of the total households were getting more than 15 ppm iodine in salt(12). Similarly, Ethiopian Demographic and Health Survey (EDHS) 2011 indicated that about a 66million persons were unprotected from iodine deficiency, and only 15% of households had access to iodized salt(8).

Although Ethiopia is enforcing the universal salt legislation to increase the content of iodine in edible salt, there is limited evidence on the level of the iodine content of salt and factors that affect its content at the household level. Thus, this study will be aimed to assess the adequacy of salt iodization, salt retailers’ knowledge, and associated factors towards iodized salt in Oromia Special Zone Surrounding Finfinne, Ethiopia.

## Methods And Materials

### Study design

A community-based cross-sectional study design was used.

### Source and study population

#### Source population

The source populations for this study were all salt retails shops and retailers in Oromia special zone Finifinne surrounding town.

#### Study population

The study populations were all the salt retail shops and retailers which are registered and licensed in 2020/2012 E.C and shop retailers over the age of 18 years of Oromia special zone Finifine surrounding.

#### Sample size determination

For this study, the sample size was determined by using a single population proportion formula at a 95% confidence interval with a 5% margin of error. Since there is no previous study in the area the proportion of iodized salt adequacy among the retailer’s shop is considered as 50%.

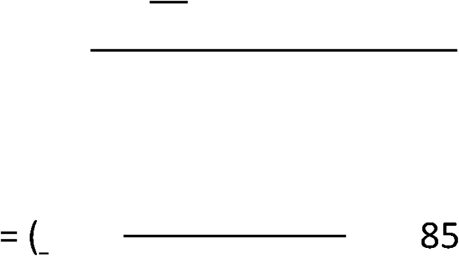

Since the study population, in the study area, is less than 10,000 a population correction formula is used.

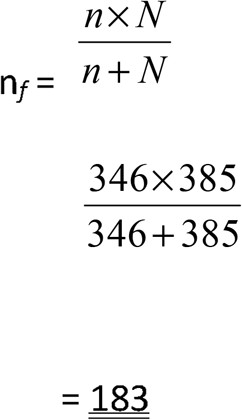

After considering the 10% non-response rate final sample size will be 202.

##### The sample size for factors associated with the outcome variables

The sample size for factors associated with the dependent variables was calculated EPI Info software version calculator 7.2.2.6 for cross-sectional study by considering the confidence interval, power, and ratio among unexposed and exposed as 95%, 80%, and 1:1 respectively.

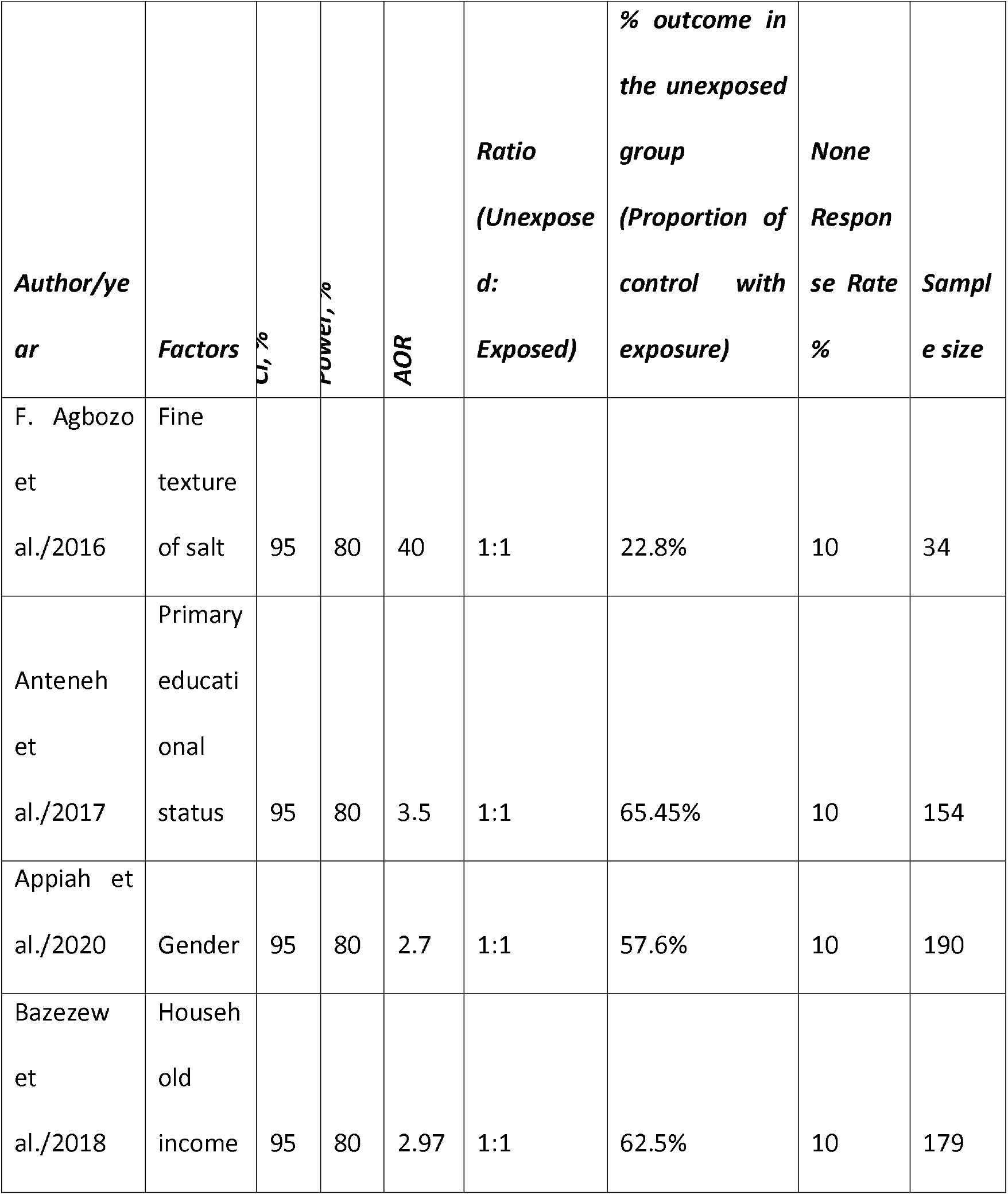

Hence, the sample size calculated for the first specific objective was greater than from the factors, we have considered it as a final sample size of the study.

##### Sampling technique

Since the study involved all the towns (six) of the Addis Ababa Surrounding Finfinne Special Zone, the calculated sample size was proportionally allocated for each town based on the calculated final sample size (202), the number of available shops in each town, and the total number of shops (346) in the Addis Ababa Surrounding Finfinne Special Zone as described below; and finally, to select the study units (retailers of salt) from each town, the researcher used simple random sampling technique.

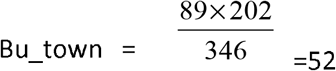

That means the sample size that was taken from Burayu town (Bu_town) is 52 shops.

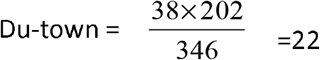

That means the sample size that was taken from Dukem town (Du_town) is 22 shops.

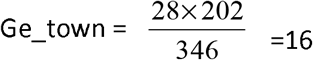

That means the sample size that was taken from Gelan town (Ge_town) is 16 shops.

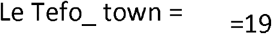

That means the sample size that was taken from Lega Tafo town (Le Tafo_town) is 19 shops.

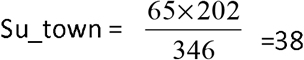

That means the sample size that was taken from Sululta town (Su_town) is 38 shops.

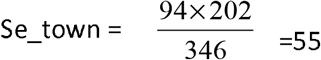

That means the sample size that was taken from Sebeta town (Se_town) is 55 shops.

##### Inclusion and exclusion criteria

###### Inclusion criteria

- All shop salt retailers who are available on the date of data collection

###### Exclusion criteria

- Shop salt retailers under the age of 18 years

###### Data collection tools and techniques

A structured questionnaire adapted from similar studies, on the iodized salt program assessment tools, (23,24,31,32) was used to conduct a face-to-face interview. Using a zipper bag, 0.5kg sample (corrosive) and 0.5 kg packed (fine granule)of salt was collected from each retail shop and the iodine level was determined using the titration method in the Ethiopian Food and Drug Authority Laboratory.

###### Procedure to test the iodine content of salt

A 10g iodized salt is dissolved in 50ml distilled water after ensuring the sample salt was thoroughly mixed in zip-lock bags. Once the salt is dissolved in the measured amount of water, 2ml sulphuric acid and 5ml potassium iodide are added to the salt solution, which in the presence of iodine will turn yellow. The reaction mixture was then kept in a dark place (with no exposure to light) for 10 min to reach the optimal reaction time before titrating with sodium thiosulfate using starch (2ml) as the direct indicator(7). Adequately iodized salt at the retail level was defined when a salt sample has 20-40 parts per million (PPM) of iodine(46), otherwise inadequate or out of specification(OOS). Eight questions assessed knowledge, retailers who scored above the mean in the overall knowledge score were considered as having good knowledge and poor otherwise.

##### Study variables

###### Dependent variables

- Adequacy of salt iodization
- Knowledge of salt retailers

###### Independent variables

- Socio-demographic characteristics of the retailer (age, sex, educational status, monthly income, and family size)
- Behavioral characteristics (access of information health information about iodized salt, media exposure)
- Controlling market salts by concerned bodies (a legal law that prohibits the selling of non-iodized salt)
- The texture of available salts in the market (fine, granular, or coarse)

###### Operational definition

If the content of iodine is 20-40PPM for iodized salt at the retail level, then it is called adequately iodized salt otherwise it’s either inadequately iodized(<20PPM, or above the standard(>40PPM)(46).

The salt shop retailers’ knowledge level was categorized as ‘good’ and ‘poor’ based on the modified Blooms cut-off points. Respondents with knowledge scores above the mean (≥ 4) were considered as good, while respondents who scored below the mean (<4) were grouped as poor.

##### Data management

###### Data quality

To maintain the quality of the data, the researcher has taken the following measures. During the designing of the study and data collection tools development, the researcher used peer-reviewed journals for the selection and development of the data collection tools, and also to have a common understanding among the respondents of the research questions which are primarily developed in the English language is translated to local language (Oromiffa). Furthermore, training for two days was given for the data collectors and supervisors focusing on the research objectives, how to conduct an interview, and how to maintain the privacy of the respondent’s information.

Before the commencement of the actual data collection, the researcher, data collectors, and supervisors conducted the pre-test on 5% of the sample size in Bishoftu town, and based on the findings the necessary modifications were applied.

During the data collection, in fieldwork, the supervisors were assisting the data collectors, controlled the completeness of the questionnaire, and if any problem happens they provided on-spot solutions to the data collectors.

After the data collection, all the questionnaires were re-checked for completeness and coded by the researcher before data entry. After data entry the missing values were cleaned by running frequency for each variable, and also the researcher checked the outliers before starting the data analysis.

### Data analysis

The collected data were coded, entered, and analyzed by using SPSS software version 23. Both descriptive and analytic statistical analysis was done. Descriptive statistics such as mean, standard deviation, and frequency distributions were performed.

After checking the model fitness by Hosmer-Lemeshow goodness-of-fit test, both bivariate and multivariate analyses were conducted to examine the association between the dependent variable and independent variables. Including the independent variables which will be statistically significant, variables with a p-value of less than 0.25 on bivariate analysis were exported to multivariable logistic analysis to control the possible confounders. The strength of association between the dependent variable and independent variables was explained by using the odds ratio and the level of statistical significance was accepted at a p-value of less than 0.05. Finally, the results were presented by using graphs and tables.

### Ethical Considerations

A formal letter of permission has been obtained from Oromia regional state Public health bureau, Health research Ethical review committee(BEFO/HR/1-16/72); After explaining the objective of the research, verbal consent was obtained from each study participant. Confidentiality of responses was maintained throughout the research process. Personal privacy and cultural norms were respected properly. No names were used; however, the questionnaires were serial numbered for data entry

## RESULTS

### Background characteristics of Retailers

A total of 202 retailers’ were approached in this study with the mean age of the retailers was 28.44 with a standard deviation of ±7.264 and over half of the study participants were in the age range of 18-28 years. Over sixty percent (125) of the participants were married.

Regarding the respondent’s monthly income, around sixty-four percent (129) of the respondents had an average monthly income of less than 5,000 Ethiopian birrs, and over half of them had a family size of 4-6. One hundred and eighty-six (92.1%) retailers were shops and the rest 3(1.5%) and 13(6.4%) were supermarkets and minimarkets, respectively (table 1).

**Table 1:** Background characteristics salt retailers.

| **Variable** | **Frequency** | **Percent** |
| --- | --- | --- |
| Sex |  |  |
| Male | 126 | 62.4 |
| Female | 76 | 37.6 |
| Age |  |  |
| 18-28 | 119 | 58.9 |
| 29-39 | 68 | 33.7 |
| >= 40 | 15 | 7.4 |
| Marital status |  |  |
| Married | 125 | 61.9 |
| Never married | 77 | 38.1 |
| Educational status |  |  |
| Read and write | 14 | 6.9 |
| Primary school | 56 | 27.7 |
| Secondary school | 96 | 47.5 |
| College diploma and above | 36 | 17.8 |
| Average monthly income |  |  |
| Less than 5000 birr | 129 | 63.9 |
| Greater than equal to 5000 birr | 73 | 36.1 |
| Family size |  |  |
| 1-3 | 77 | 38.1 |
| 4-6 | 109 | 54.0 |
| >= 7 | 16 | 7.9 |
| Type of shop |  |  |
| Supermarket | 3 | 1.5 |
| Minimarket | 13 | 6.4 |
| Shop | 186 | 92.1 |
| Ethiopian law bans selling non-iodized salt |  |  |
| Yes | 57 | 28.2 |
| No | 145 | 71.8 |

#### Iodine concentration levels of salts in the Retailers shop

The mean iodine level of the salts was 32.16 PPM (parts per million) with a standard deviation of ± 14.337, around 32(19.3%) retail shops were selling inadequately iodized salt, and also 39(23.5%) were selling salts that are iodized above the standard.

More than half (57.2%) of salts in retail shops in the districts have satisfactory (20-40 PPM) iodine levels (figure 3), around 42.8% (71) have inadequate or out of the specification iodine concentration level.

**Figure 1:**
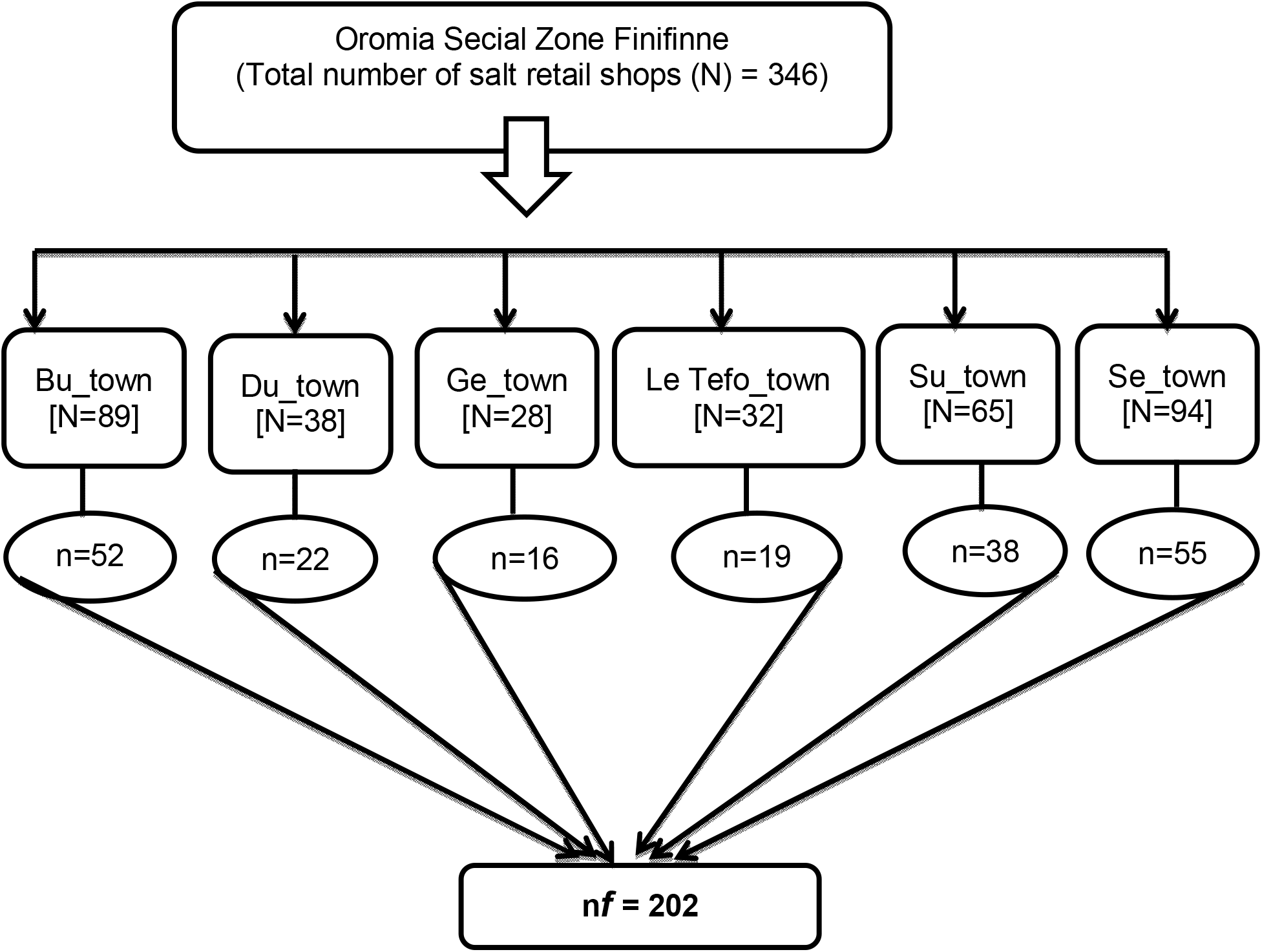
Schematic presentation of sampling technique for the assessment of the adequacy of salt iodization, knowledge of retailers, and associated factors about iodized salt in **Addis Ababa Surrounding Finfinne Special Zone** Oromia regional state, Ethiopia.

**Figure 2:**
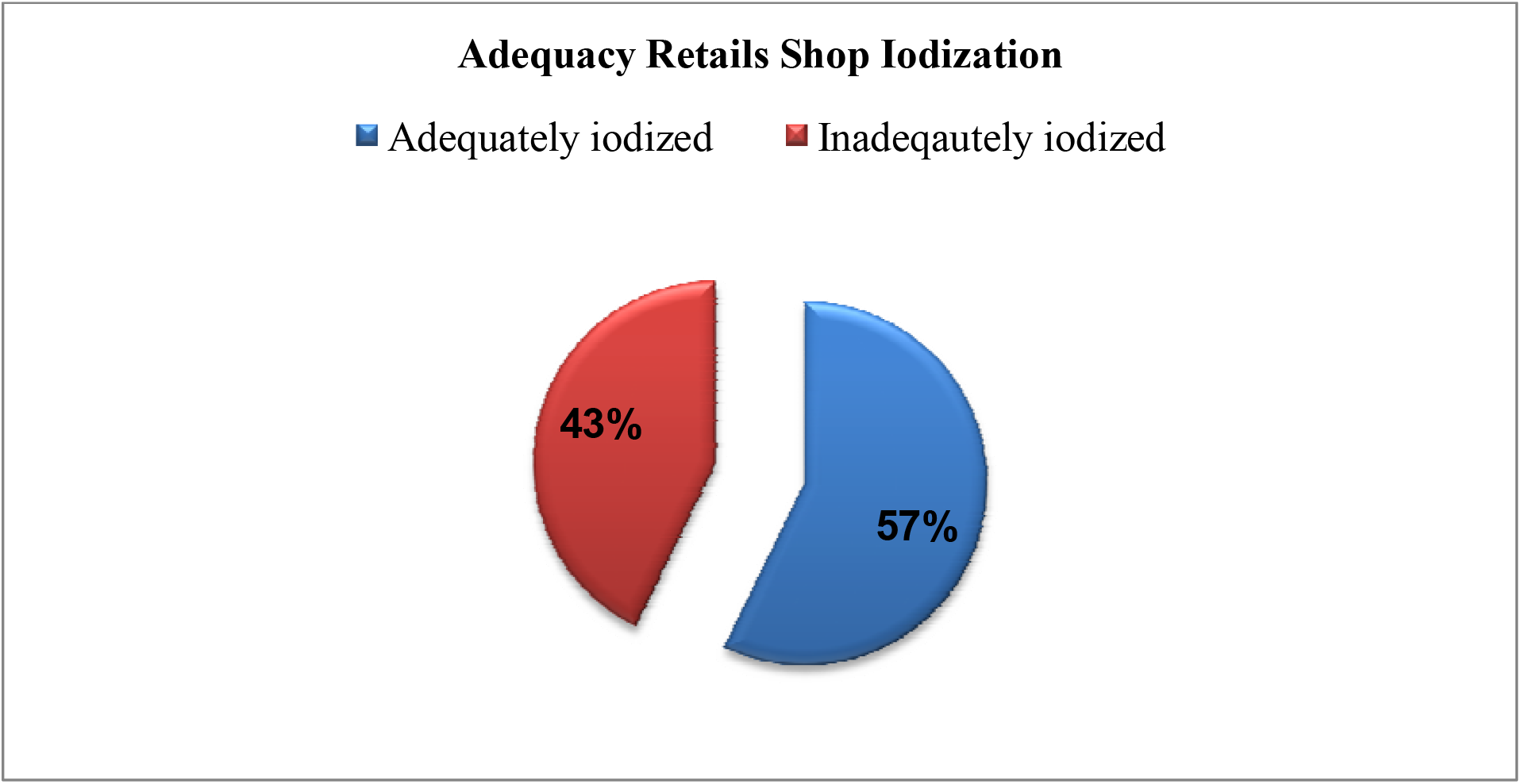
Adequacy of salt iodization among retailer shops in Addis Ababa Surrounding Finfinne Special Zone Ethiopia.

**Figure 3:**
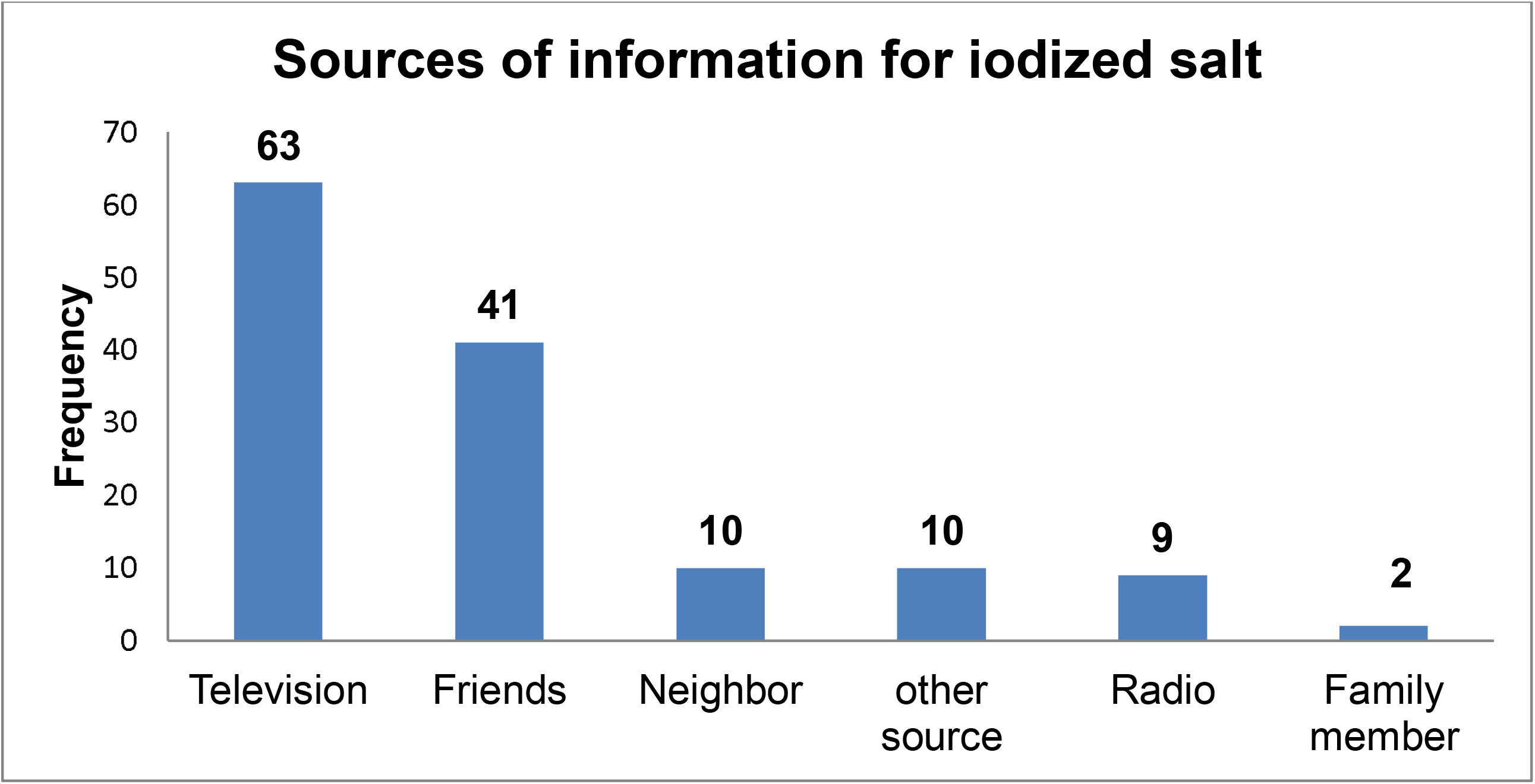
Source of information about iodized salt among the salt retailer in Addis Ababa surrounding Finfinne Special Zone, 2021.

#### The texture of salt type, how it was displayed for sale, and adequacy of iodized salt

Fine-texture 64(57.7%) and coarse-texture 15(57.7%) have adequate iodine (20-40 ppm). Salt kept in the single package from the producer (41.4%), in a double bag 3(75%), and in an open container 1(33.3%), placed under a shade 70 (42.9%), and placed directly in the sun 0(0.0%) have inadequate iodine (<20 ppm). Also, salt kept in an open container 2(66.7%), placed under a shade 78(58.6%), and placed directly in the sun 10(100.0%) have adequate iodine (20-40 ppm).

Twenty-six (37.7%) and over 46% of retailers with poor and good knowledge levels were selling inadequate iodine salt (<20 ppm) in their shop during the study period respectively (Table 2).

**Table 2:** Texture of salt type, how it was displayed for sale, retailers’ knowledge and adequacy of iodized salt.

| **Variable** | **Adequately Iodized** | **Inadequately Iodized** | **Total** |
| --- | --- | --- | --- |
| The texture of salt in the shop | n (%) | n (%) | n |
| Fine granule | 64(57.7) | 47(42.3) | 111 |
| Coarse granule | 15(57.7) | 11(42.3) | 26 |
| How to store |  |  |  |
| With closed container | 93(57.1) | 70(42.9) | 163 |
| With open container | 2(66.7) | 1(33.3) | 3 |
| Type of package |  |  |  |
| Primary/single | 78(58.6) | 55(41.4) | 133 |
| Secondary/double | 1(25.0) | 3(75.0) | 4 |
| Knowledge level about iodized salt |  |  |  |
| Good knowledge | 52(53.6) | 45(46.4) | 97 |
| Poor knowledge | 43(62.3) | 26(37.7) | 69 |
| Expose salt to sunlight |  |  |  |
| Yes | 10(100) | 0(0.0) | 10 |
| No | 85(54.5) | 71(42.8) | 156 |

#### Factors associated with adequacy of salt iodization

To determine factors associated with the adequacy of salt iodization binary bivariable analysis was performed for socio-demographic and other variables. Based on this analysis educational status of the respondents had a significant association with the adequacy of salt iodization (Table 3).

**Table 3:** Factors associated with the adequacy of salt iodization.

| Variables | Salt Iodization |  | COR | P-value | 95% CI |
| --- | --- | --- | --- | --- | --- |
|  | Adequately | Inadequately |  |  |  |
|  | Iodized | Iodized |  |  |  |
| Sex |  |  |  |  |  |
| Male | 60 | 46 | 1.07 | 0.829 | 0.56-2.036 |
| Female | 35 | 25 | 1:00 |  |  |
| Marital status |  |  |  |  |  |
| Married | 57 | 44 | 1:00 |  |  |
| Never married | 38 | 27 | 0.920 | 0.797 | 0.49-1.73 |
| Educational level |  |  |  |  |  |
| Read and write | 7 | 3 | 0.248 | 0.077 | 0.053-1.16 |
| Primary school | 31 | 16 | 0.299 | 0.013 | 0.11-0.77 |
| Secondary school | 46 | 33 | 0.415 | 0.047 | 0.17-0.98 |
| College diploma and above | 11 | 19 | 1:00 |  |  |
| Monthly income |  |  |  |  |  |
| Less than 5000 | 65 | 40 | 0.596 | 0.111 | 0.31-1.12 |
| ≥ 5000 | 30 | 31 | 1:00 |  |  |
| Family size |  |  |  |  |  |
| 1-3 | 34 | 27 | 1:00 |  |  |
| 4-6 | 53 | 39 | 0.927 | 0.819 | 0.48-1.78 |
| ≥7 | 8 | 5 | 0.787 | 0.702 | 0.23-2.68 |
| Knowledge about iodized salt |  |  |  |  |  |
| Good | 52 | 45 | 1:00 |  |  |
| Poor | 43 | 26 | 0.699 | 0.26 | 0.37-1.311 |
| How to store to sale |  |  |  |  |  |
| With closed container | 93 | 70 | 1:00 |  |  |
| With open container | 2 | 1 | 1.39 | 0.814 | 0.086-22.72 |
| Type of package |  |  |  |  |  |
| Primary/Single | 78 | 55 | 1:00 |  |  |
| Secondary/Double | 1 | 3 | 0.66 | 0.74 | 0.05-7.47 |
| Texture of salt |  |  |  |  |  |
| Fine granule | 64 | 47 | 1:00 |  |  |
| Coarse granule | 15 | 11 | 0.999 | 0.997 | 0.42-2.37 |

#### Multivariable binary logistic regression analysis for factors associated with inadequacy of iodized salt

To control the potential confounder’s variables with a p-value less than 0.25 in bivariable analysis such as monthly income and educational status were again analyzed in multivariable analysis. Based on this analysis of the educational status, those who were at primary school (AOR=0.32, CI: 0.12-0.8, p=0.024) levels were associated with the selling of inadequately iodized salt. Retailers who were at the primary school level were 68% (AOR=0.32, CI: 0.12-0.8, p=0.0024) were less likely to have poor knowledge than those who were at college diploma and above (table 4).

**Table 4:**
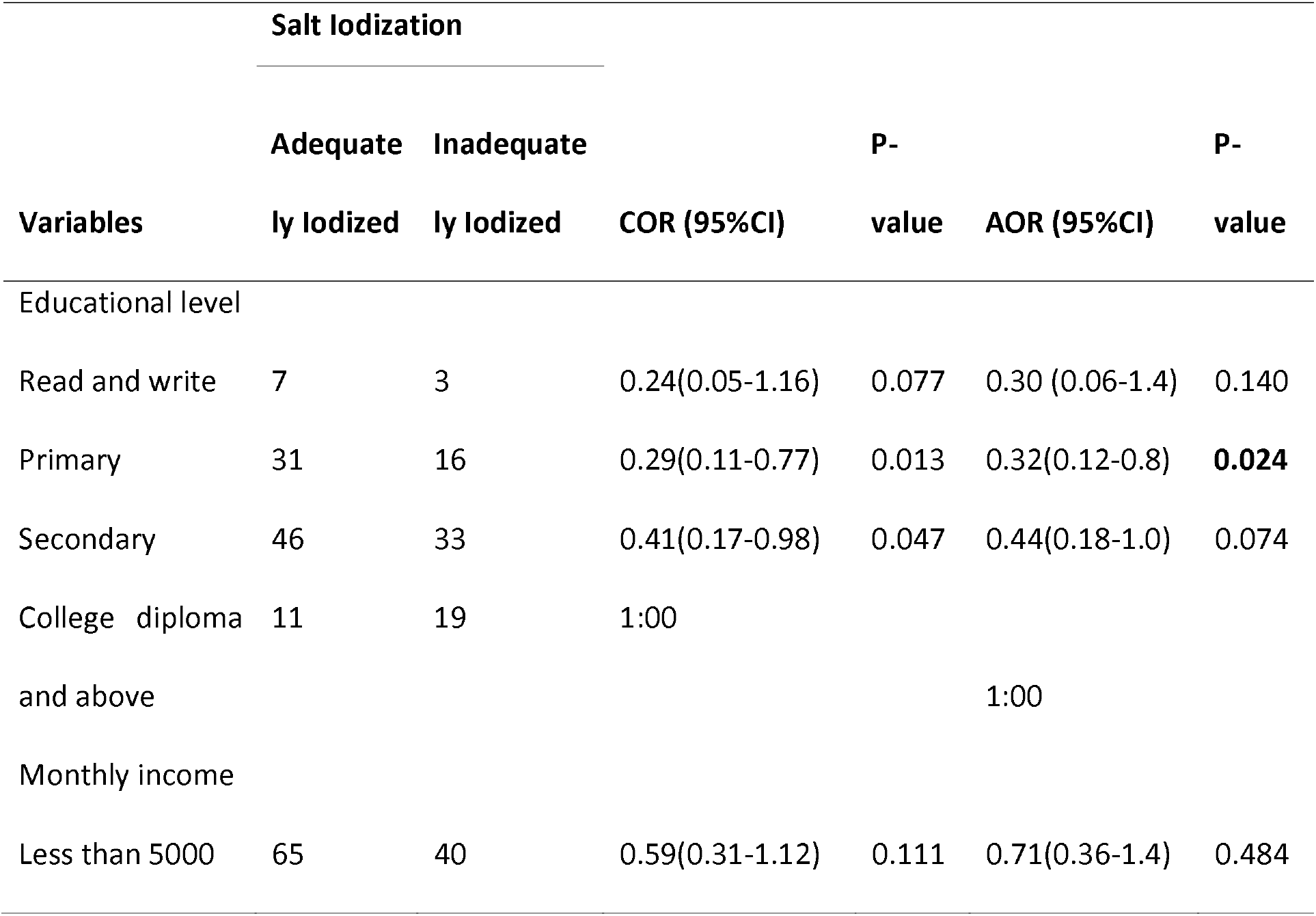
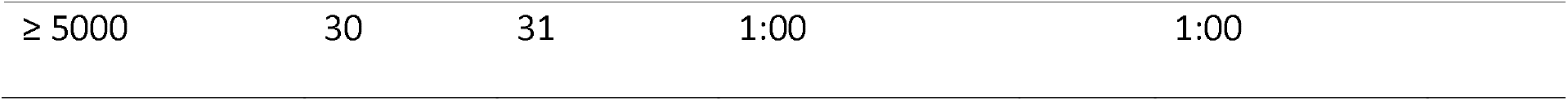
Factors associated with the adequacy of salt iodization.

#### Retailers’ primary information and knowledge of iodized salt

Of the 202 iodized salt retailers, 135(66.8%) of them heard about iodized salt and their primary source of information about iodized salt among the retailer was 63 (46.7%) television followed by friends 41 (30.4%), and none of the salt retailers got information from the health workers (Figure 4).

**Figure 4:**
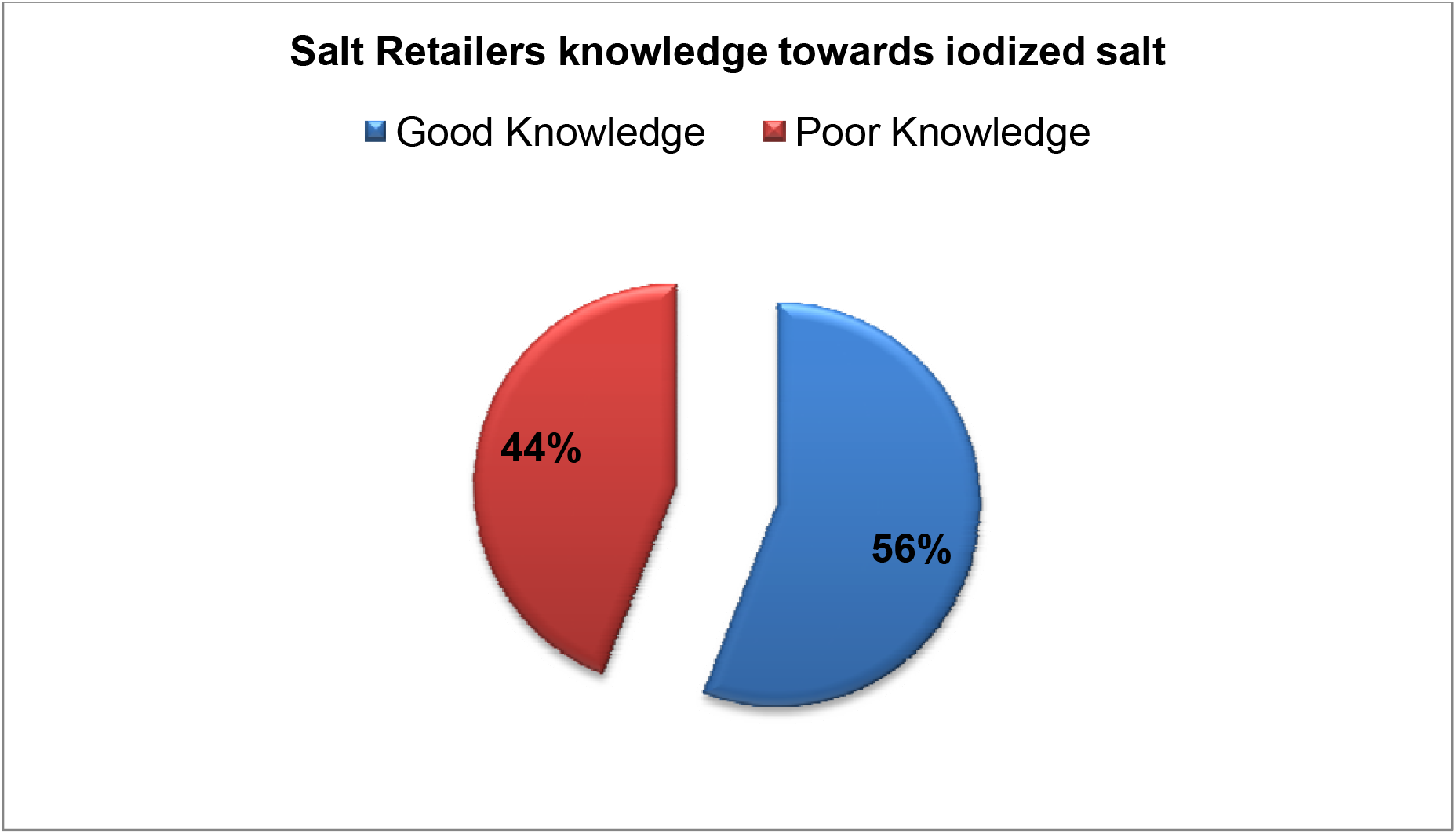
Knowledge on iodized salt among the salt retailer in Addis Ababa surrounding Finfinne Special Zone.

The mean knowledge level of the retailers towards iodized salt was found to be 4.41±2.166, and around 55.9 %(113) retailers had a good knowledge level (Figure 5). Around sixty-one percent (123) of the salt retailer knew the importance of iodized salt; 74(60.2%), 3(2.4%), 35(28.5%), and 11(8.9%) of them described that iodized salt is important for the prevention of goiter, growth, and development, for health and another purpose respectively. Over 60 % (122) of the salt retailers described different types of iodine deficiency-related health consequences; around forty-seven percent (95) of them mentioned goiter and 10 (8.2%) mental retardation as an iodine deficiency-related disorders.

Of the 202 salt retailers, only 47 (23.3%) respondents knew that iodized salt is the richest source of iodine and over two-thirds (135) of them didn’t know the source of iodine (Table 5).

**Table 5:** **Knowledge of the respondents towards iodized salt in Addis Ababa surrounding Finfinne Special Zone Ethiopia 2021**

| **Variable** | **Frequency** | **Percent** |
| --- | --- | --- |
| Heard about iodized salt |  |  |
| Yes | 135 | 66.8 |
| No | 67 | 33.2 |
| Knew the importance of iodized salt |  |  |
| Yes | 123 | 60.9 |
| No | 79 | 39.1 |
| Importance of iodized salt |  |  |
| Prevention of goiter | 74 | 60.2 |
| Growth and development | 3 | 2.4 |
| For health | 35 | 28.5 |
| For another purpose | 11 | 8.9 |
| Richest sources of iodine |  |  |
| Milk and milk products | 2 | 1.0 |
| Iodized salt | 47 | 23.3 |
| Other | 18 | 8.9 |
| I don't know | 135 | 66.8 |
| Knew the consequences of iodine deficiency |  |  |
| Yes | 122 | 60.4 |
| No | 80 | 39.6 |
| Disorders related to iodine deficiency |  |  |
| Mental retardation | 10 | 8.2 |
| Goiter | 95 | 77.9 |
| Other | 17 | 8.4 |
| Expose salt to sunlight |  |  |
| Yes | 11 | 5.4 |
| No | 191 | 94.6 |
| How do you store the salt |  |  |
| In closed container | 199 | 98.5 |
| In open container | 3 | 1.5 |

#### Factors associated with retailers’ knowledge of iodized salt

To determine factors associated with the knowledge of iodized salt both bivariable and multivariable binary logistic regression analysis were performed.

On bivariate analysis marital status being never married (p=0.01), those who heard about iodized salt (p<0.001), and who knew the availability of legal law that prohibit selling of non-iodized salt (p<0.001) were associated with having good knowledge towards iodized salt (table 6).

**Table 6:** Factors associated with retailers’ knowledge of iodized salt.

| Attribute | Knowledge level |  | COR(95%CI) | P-value | AOR(95% CI) | P-value |
| --- | --- | --- | --- | --- | --- | --- |
|  | Good | Poor |  |  |  |  |
| Sex |  |  |  |  |  |  |
| Male | 65 | 61 | 1:00 |  |  |  |
| Female | 48 | 28 | 1.60(0.89-2.88) | 0.110 | 1.5(0.66-3.5) | 0.318 |
| Age |  |  |  |  |  |  |
| 18-28 | 60 | 59 | 1:00 |  | 1:00 |  |
| 29-39 | 44 | 24 | 1.8(0.97-3.33) | 0.060 | 1.05(0.40-2.72) | 0.92 |
| >=40 | 9 | 6 | 1.47(0.49-4.40) | 0.486 | 2.50(0.51-12.07) | 0.25 |
| Marital status |  |  |  |  |  |  |
| Married | 79 | 46 | 1:00 |  | 1:00 |  |
| Never married | 34 | 43 | 0.46(0.25-0.82) | 0.009 | 0.83(0.33-2.1) | 0.73 |
| Educational level |  |  |  |  |  |  |
| Read and write | 4 | 10 | 0.15(0.039-0.60) | 0.007 | 0.54(0.08-3.6.37) | 0.52 |
| Primary school | 25 | 31 | 0.31(0.12-0.76) | 0.011 | 0.42(0.13-1.27) | 0.125 |
| Secondary school | 58 | 38 | 0.58(0.25-1.35) | 0.212 | 1.3(0.44-3.85) | 0.62 |
| College diploma and above | 26 | 10 | 1:00 |  |  |  |
| Monthly income |  |  |  |  |  |  |
| Less than 5000 | 71 | 58 | 0.90(0.50-1.61) | 0.731 |  |  |
| ≥ 5000 | 42 | 31 | 1:00 |  |  |  |
| How to store salt |  |  |  |  |  |  |
| With closed container | 87 | 112 | 1:00 |  | 1:00 |  |
| With open container | 1 | 2 | 0.38(0.035-4.35) | 0.44 |  |  |
| Heard of iodized |  |  |  |  |  |  |
| salt |  |  |  |  |  |  |
| Yes | 104 | 31 | 1:00 |  | 1:00 |  |
| No | 9 | 58 | 0.046(0.02-0.10) | **<0.001** | 0.043(0.017-0.109) | **<0.001** |
| Do you know legal |  |  |  |  |  |  |
| laws bans the |  |  |  |  |  |  |
| selling of non- |  |  |  |  |  |  |
| iodized salt |  |  |  |  |  |  |
| Yes | 44 | 13 | 1:00 |  | 1:00 |  |
| No | 69 | 76 | 0.26(0.133-0.54) | **<0.001** | 0.24(0.098-0.61) | **0.003** |

To control the potential confounders, multivariable binary logistic regression analyses were carried out for variables with a p-value less than 0.25 on bivariate analysis. Based on the multivariable binary logistic regression analysis those who heard about iodized salt and having legal law that bans selling non-iodized salt were associated with good knowledge among the salt retailers.

Retailers who didn’t hear about iodized salt were 95.7% (AOR=0.043: CI, 0.017-0.109, p<0.001) times less likely to have poor knowledge than those who heard about iodized salt, and retailers who didn’t know the availability of legal law prohibits the selling of non-iodized salt were 76% (AOR=0.24, CI: 0.098-0.61, p=0.029) times more likely to have poor knowledge than who did know.

## DISCUSSION

One of the most common and effective public health approaches for the eradication of IDD globally is universal salt Iodization(29). The Ethiopian Council of Ministers passed salt legislation in March 2011; according to this regulation, every salt for human consumption needs to be iodized, and any iodized salt for human consumption shall conform to the standards for iodized salt set by the appropriate authority(8).

In this study the retailers in the Addis Ababa surrounding Finfinne Special Zone 56% have good knowledge of iodized salt and total retailers’ salt samples, 57.2% were adequately (20-40ppm) iodized. The number of retail shops trading adequately iodized (20-40ppm) salt in the districts was not surprising. Nevertheless, the result is above what was reported from surveys EDHS(4,17,31,32) and the result is below the findings of Uganda (50) and Saudi Arabia (24). Although the iodine concentration level of salt in retail shops in the district may be above other parts in Ethiopia, it is still scantily above half of the 90% target suggested by the WHO/UNICEF(51). These differences might be due to challenges in the distribution chain and require policies and laws to promote the production and sale of iodized salt to be revisited and strengthened.

The primary source of information on iodized salt could affect an individual’s knowledge on iodized salt and utilization of iodized salt(31,49). This study showed that just 48.7% of salt retailers got information about iodized salt from television; this agrees with Bazezew et al.’s study in Addis Ababa, Ethiopia(48). In this study, none of the salt retailers got information about the iodized salt from the health care workers. However, this finding disagrees with the study in Ghana(31).

The district has more educated males than females(45). Therefore; it is not unusual that females were less likely to have high knowledge of iodized salt than males. The finding agrees with studies done in the northern parts of Ethiopia that indicated gender inequalities in education(43,48,49).

This study showed that retailers who placed salt under shade were more likely to have high knowledge of iodized salt than retailers who placed salt directly in the sun. However, direct exposure to iodized salt to the sun is known to reduce its iodine content(48). The practice of placing salt directly in the sun could be avoided if the individual knows the effects of exposing iodized salt to the sun’s rays. Again, this study showed that retailers who attained basic, secondary, and tertiary education were more likely to have high knowledge of iodized salt than retailers who read and write. Also, retailers who got information about iodized salt from television were more likely to have good knowledge of iodized salt than retailers who got information about iodized salt from other sources. What these findings mean is that the quality of the information received and an improvement in an individual’s education can influence his/ her knowledge of iodized salt. These findings agree with other studies elsewhere(31,43,48,49).

Furthermore, this study exposed the significant association between iodine content/adequacy of salt iodization and educational status of the retailers, where retailers who are over the educational level of primary and above were very well aware of the importance of iodized salt, how to store and benefit of selling them(4,33,48).

A study from Ghana revealed a significant association between adequacy of salt iodization and sunlight exposure(31). However, the study didn’t report a significant association between the adequacy of salt iodization and sunlight exposure. This discrepancy may be due to the difference in sample size.

## Data Availability

All relevant data are within the manuscript and its Supporting Information files.

## Acknowledgment

Authors please Oromia regional state Public health emergency research office; for their cooperation and all participants.

## Authors Contributions

All authors made notable pieces from development to interpretation of data, contributed in from drafting to amend and gave final approval of the version to be published; and agree to be responsible for all features of this paper.

## Conclusion

This study revealed that lower adequacy of salt iodization at salt retailers’ shop level compared to from the global target point. Educational level was significantly associated with the adequacy of salt iodization.

Over half of the salt retailers had good knowledge of iodized salt. Salt retailers who heard of iodized salt and having legal framework/law that prohibits the selling of non-iodized salt were significantly associated with the good knowledge among the retailers.

## ABBREVIATIONS

AIS: Adequately Iodized Salt
CES: Compulsory Ethiopian Standard
CSA: Central Statistical Agency
EDHS: Ethiopian Demographic and Health Survey
ID: Iodine Deficiency
IDD: Iodine Deficiency Disorders
IQ: Intelligence Quotient
IRB: Institutional Review Board
PPM: Parts Per Millions
PI: Principal Investigator
RTK: Rapid Test Kits
USI: Universal Salt Iodization
UNICEF: United Nations International Children’s Emergency Fund
WHO: World Health Organization

## REFERENCES

1. Mekonnen TC, Eshete S, Wasihun Y, Arefaynie M, Cherie N. Availability of adequately iodized salt at household level in Dessie and Combolcha Towns, South Wollo, Ethiopia. BMC Public Health. 2018;18(1):1–9.

2. Tariku WB, Mazengia AL. Knowledge and Utilization of Iodized Salt and Its Associated Factors at Household Level in Mecha district, Northwest Ethiopia. J Nutr Metab. 2019;2019.

3. Kapil U. Health Consequences of Iodine Deficiency. 2007;7(3):267–72.

4. BG, KA, A A, M K, Y TY, HR S. Availability of Adequate Iodized Salt at Household Level and Associated Factors in Rural Communities in Laelay Maychew District, Northern Ethiopia: A Cross Sectional Study. J Nutr Heal Sci. 2015;1(4):1–9.

5. Mannar MGV, Bohac L. Achieving Universal salt iodization: lesson learned and emerging issues. 2008.

6. Bauer A. Iodine Deficiency [Internet]. Vol. 2, British Medical Journal. 2019. p. 831. Available from: www.thyroid.org

7. World Health Organization. Assessment of iodine deficiency disorders and monitoring their elimination. A guide for programme managers. 2007.

8. Federal Democratic Republic of Ethiopia. Federal Negarit Gazeta of the Federal Democratic Republic of Ethiopia. Council of Ministrers Regulation No. 204/2011, Addis Ababa. Federal Negarit Gazeta [Internet]. 2011;5785. Available from: https://www.google.com/search?q=International.+CSAEaI.+Ethiopia+demographic+and+HealthSurvey+2011.+Addis+Ababa%3A+Ethiopia+Central+Statistical+Agency+and+ICF+International%3B+2012.&sxsrf=ALeKk03FuHha8245wGMzb7aXbXdDRZFLcw%3A1619854564403&ei=5ASNYKn_F8eGhb

9. GlobalIodinenetwork. REPORT ON THE 2019 CONSULTATION ON Sustainable Prevention and Control of Iodine Deficiency Disorders in Eastern and Southern Africa. Dares Salaam, Nairobi, Seattle; 2020.

10. Development Initiatives. Global Nutrition Report:Shining a light to spur action on nutrition. Bristol, UK: Development Initiatives Poverty Research Ltd; 2018.

11. Andersson M, Karumbunathan V, Zimmermann MB. Global iodine status in 2011 and trends over the past decade. J Nutr. 2012;142(4):744–50.

12. Ethiopian Public Health Institute. Ethiopian National Micronutrient Survey report [Internet]. Addis Ababa, Ethiopia; 2016. Available from: https://www.google.com/search?q=Ethiopian+Public+Health+Institute.+National+Micronutrient+Survey+Report+Addis+Ababa%3A+Federal+Ministry+of+Health%2C+2016+2016&sxsrf=ALeKk01YLkDj6YK6KR61L-wPBbPdAuLSbw%3A1619854411938&ei=SwSNYLToOIv0gQaL8aroDA&oq=Ethiopian+

13. United Nations International Children’s Emergency Fund. Sustainable Elimination of Iodine Deficiency: Progress since the 1990 World Summit for Children. Geneva: The United Nations International Children’s Emergency Fund; 2006.

14. KA A, BT A, AM G, T Y, Z A, Abuye C. Iodine Deficiency disorders in Burie and Wombwema district, West Gojjam, Ethiopia. African J Food, Agric Nutr Dev.4806;4(Idd):9167–81.

15. Hailu S, Wubshet M, Woldie H, Tariku A. Iodine deficiency and associated factors among school childrenllJ: a cross-sectional study in Ethiopia. Arch Public Heal [Internet]. 2016;74(46):1–7. Available from: http://dx.doi.org/10.1186/s13690-016-0158-4

16. Muktar M, Teji Roba K, Mengistie B, Gebremichael B. Iodine deficiency and its associated factors among primary school children in Anchar district, Eastern Ethiopia. Pediatr Heal Med Ther. 2018;9:89–95.

17. Ethiopia Demographic and Health Survey. Central Statistical Agency[Ethiopia] and ICF [Internet]. Addia Ababa, Rockville, Maryland, Ethiopia, USA: CSA and ICF; 2016. Available from: https://www.google.com/search?q=International.+CSAEaI.+Ethiopia+demographic+and+HealthSurvey+2011.+Addis+Ababa%3A+Ethiopia+Central+Statistical+Agency+and+ICF+International%3B+2012.&sxsrf=ALeKk03FuHha8245wGMzb7aXbXdDRZFLcw%3A1619854564403&ei=5ASNYKn_F8eGhb

18. World Health Organization. Global prevalence of iodine deficiency disorders Micronutrient deficiency information system (MDIS) working paper no. 1 [Internet]. 2004. 80 p. Available from: http://www.who.int/nutrition/publications/micronutrients/iodine_deficiency/54015_mdis_workingpaper1/en/

19. Pearce EN, Lazarus JH, Moreno-Reyes R, Zimmermann MB. Consequences of iodine deficiency and excess in pregnant women: an overview of current knowns and unknowns. Am J Clin Nutr. 2016;104:918S–923S.

20. Asfaw A, Belachew T. Magnitude of iodine deficiency disorder and associated factors in Dawro zone, Southwest Ethiopia; the hidden hunger: A cross-sectional study. BMC Nutr. 2020;6(1):1–10.

21. World Health Organization. Guideline: Fortification of food-grade salt with iodine for the prevention and control of iodine deficiency disorders. Who Guideline. Geneva: World Health Organization; 2014.

22. Abebe Z, Tariku A, Gebeye E. Availability of adequately iodized in Northwest Ethiopia: A cross-sectional study. Arch Public Heal. 2017;75(1):1–9.

23. Agbozo F, Der JB, Glover NJ, Ellahi B. Household and market survey on availability of adequately iodized salt in the Volta region, Ghana. Int J Heal Promot Educ [Internet]. 2017;55(3):110–22. Available from: http://dx.doi.org/10.1080/14635240.2016.1250658

24. Al-Dakheel MH, Haridi HK, Al-Bashir BM, Al-Shangiti AM, Al-Shehri SN, Hussein I. Assessment of household use of iodized salt and adequacy of salt iodization: A cross-sectional National Study in Saudi Arabia. Nutr J. 2018;17(1):1–7.

25. World Health Organization. Iodine status worldwide. WHO Glob Database Iodine Defic. 2004;1–12.

26. World Health Organization Regional Committee for South-East Asia. WHO Regional Committee for South-East Asia. South Asia Iodine Glob Netw. 2012;(September):8–9.

27. Iodine Global Network. Global Scorecard of iodine nutrition in 2020: optimal iodine intake in 131 countries. IDD Newsl. 2020;(26):1–14.

28. Iodineglobalnetwork. Iodine Global Network; Annual Report. 2019.

29. Jain S, Kilyani M, Walters D, Khan N. The Need for Investment in Universal Salt Iodization. Nutrition International: Nourish life. 2020.

30. Venkaktesh Mannar MG. Making salt iodization truly universal by 2020. Iodine Glob Netw Newsl. 2014;(May 2014):12–5.

31. Appiah PK, Fenu GA, Yankey FWM. Iodine Content of Salt in Retail Shops and Retailers’ Knowledge on Iodized Salt in Wa East District, Upper West Region, Ghana. J Food Qual. 2020;2020.

32. Afework A, Mulu W, Abate A, Lule A. Handling and Adequacy of Iodine at Household Level: Community Based Cross-sectional Survey in Dega Damot District, West Gojjam Zone, Amhara Regional State, Ethiopia. bioRxiv. 2019;

33. Anteneh ZA, Engidayehu M, Abeje G. Iodine content of dietary salt at household level and associated factors using Iodometric titration methods in Dera District, Northwest Ethiopia. BMC Nutr. 2017;3(1):1–7.

34. Berhane A, Baraki N, Endale BS. Availability of Adequately Iodized Salt at Household Level and Associated Factors in Dire Dawa, Eastern. 2016;5(4):392–9.

35. Regassa D M, H Tw. Utilization of Adequately Iodized Salt on Prevention of Iodine Deficiency Disorders at Household Level and Associated Factors in Lalo Assabi District, West Ethiopia. J Nutr Food Sci. 2016;06(02).

36. SB H, S L. Proper Utilization of Adequatly Iodized Salt at House Hold Level and Associated Factores in Asella Town Arsi Zone Ethiopia: A Community based Cross Sectional Study. J Food Process Technol. 2016;07(04).

37. Desta AA, Kulkarni U, Abraha K, Worku S, Sahle BW. Iodine level concentration, coverage of adequately iodized salt consumption and factors affecting proper iodized salt utilization among households in North Ethiopia: A community based cross sectional study. BMC Nutr. 2019;5(1):1–10.

38. Dinka AW, Kebebe T, Nega G. Iodine Level of Salt and Associated Factors at Household Level in Gidami District, Oromia Region, Ethiopia: A Cross-Sectional Study. Nutr Diet Suppl. 2021;Volume 13:9–16.

39. Dida N, Legese A, Aman A, Muhamed B, Damise T, Birhanu T, et al. Availability of adequately iodised salt at household level and its associated factors in Robe town, Bale Zone, South East Ethiopia⍰: community-based cross-sectional study Availability of adequately iodised salt at household level and its associated fac. South Africa Joural Clin Nutr [Internet]. 2020;58–63. Available from: https://doi.org/10.1080/16070658.2018.1551767

40. Ajema D, Bekele M, Yihune M, Tadesse H, Gebremichael G. Socio ⍰ demographic correlates of availability of adequate iodine in household salt⍰: a community ⍰ based cross ⍰ sectional study. BMC Res Notes [Internet]. 2020;125(13):1–6. Available from: https://doi.org/10.1186/s13104-020-04983-w

41. Yazew T. Availability of Adequately Iodized Salt at Household Level and Its Associated Factors in Horro Woreda, Horro Guduru Wollega Zone, Oromia, Ethiopia. Pathol Lab Med. 2020;4(1):20.

42. Tsegaye M, Hailu D, Zegeye M. Availability and Utilization of Adequately Iodized Salt by Urban and Rural Households and Associated Factors in Southern Ethiopia, Sidama Zone, Bensa Woreda⍰: A Comparative Cross-sectional Study. Int J Food Sci Nutr Eng. 2016;6(3):69–71.

43. Dessu S, Dawit Z, Alemu G. Assessment of Knowledge on Iodized Salt Utilization and Associated Factors among Households in Arba Minch Town, Southern Ethiopia. Int J Res Stud Med Heal Sci. 2018;3(12):1.

44. Yeshaw Y, Alem AZ, Tesema GA, Teshale AB, Liyew AM, Tesema AK. Spatial distribution and determinants of household iodized salt utilization in Ethiopia: a spatial and multilevel analysis of Ethiopian demographic and Health survey. BMC Public Health. 2020;20(1):1–18.

45. Terfa BK, Chen N, Zhang X, Niyogi D. Urbanization in small cities and their significant implications on landscape structures: The case in Ethiopia. Sustain. 2020;12(3):1–19.

46. Ethiopian Standards Agency. Compulsory Ethiopian Standard Iodized edible salt Specification [Internet]. Addis Ababa; 2017. (CES 70). Report No.: ICS:67.220.20. Available from: https://www.nrc.no/globalassets/pdf/tenders/ethiopia/dec-2020-procurement-of-different-emergency-food-and-nonfood-items/annex-1_ces-70-ethiopian-standard.pdf

47. Egziabher TBG, Edwards S. Proper Utilization of Iodized Salt and assoaciated factors among rural community of hetosa district, Oromia Regional State, southeast Ethiopia. Africa’s potential Ecol Intensif Agric. 2013;53(9):1689–99.

48. Bazezew MM, Yallew WW, Belew AK. Knowledge and practice of iodized salt utilization among reproductive women in Addis Ababa City. BMC Res Notes [Internet]. 2018;11(1):1–8. Available from: https://doi.org/10.1186/s13104-018-3847-y

49. Senbeta AM, Mamo FT, Desalegn BB, Daba AK. Knowledge and practices of iodized salt utilization, health consequences, and iodine concentration on dietary salts at retailer and households in Jigjiga town, Somali, Ethiopia. Cogent Food Agric [Internet]. 2021;7(1). Available from: https://doi.org/10.1080/23311932.2021.1911421

50. Knowles JM, Garrett GS, Gorstein J, Kupka R, Situma R, Yadav K, et al. Household coverage with adequately iodized salt varies greatly between countries and by residence type and socioeconomic status within countries: Results from 10 national coverage surveys. J Nutr. 2017;147(5):1004S–1014S.

51. United Nations Children’s Fund. The State of the World’s Children: A fair chance for every child [Internet]. 2016. Available from: https://www.google.com/url?sa=t&rct=j&q=&esrc=s&source=web&cd=&cad=rja&uact=8&ved=2ahUKEwjEzZTx7s3zAhWDoFwKHa56DhwQFnoECA8QAQ&url=https%3A%2F%2Fwedocs.unep.org%2Fbitstream%2Fhandle%2F20.500.11822%2F7531%2F-The_state_of_the_worlds_children_2016_A_fair_chan

